# Uptake and predictors of viral load testing and viral suppression among people receiving antiretroviral therapy in mainland Tanzania

**DOI:** 10.64898/2026.03.11.26348142

**Authors:** Ritah F. Mutagonda, Meshack D. Lugoba, James Mwakyomo, Veryeh Sambu, George Musiba, Beatrice Mutayoba, Mercy Mpatwa, Prosper Njau, Werner Maokola, Raphael Z. Sangeda

## Abstract

**Background:** Routine viral load monitoring is essential for assessing treatment effectiveness and for achieving global HIV treatment targets. Although viral load testing capacity has expanded across sub-Saharan Africa, gaps remain in monitoring coverage and equitable treatment outcomes. National-level analyses examining viral load uptake, suppression, and geographic variation in Tanzania remain limited.

**Methods:** We conducted a retrospective observational analysis using routinely collected HIV programme data from the National AIDS and Sexually Transmitted Infections Control Programme (NASHCoP) Care and Treatment Clinic database. The analysis included 70,000 individuals receiving antiretroviral therapy between 2017 and 2021 across all regions of mainland Tanzania. Viral load uptake was defined as the presence of at least one documented viral load test during the observation period. Viral suppression was defined as a viral load result <1,000 copies/mL among individuals with available test results. Descriptive analyses summarized demographic and treatment characteristics, while multivariable logistic regression models identified predictors of viral load uptake and viral suppression. Geographic variation in programme performance was examined at regional and district levels using choropleth maps.

**Results:** Among 70,000 individuals included in the analysis, 49,820 (71.2%) had at least one documented viral load result. Among those tested, 44,187 (88.7%) achieved viral suppression. Viral load uptake was lowest among young adults and slightly lower among males than females. Viral suppression was lower among children and adolescents than among older age groups. Individuals receiving dolutegravir-based regimens demonstrated higher viral suppression than those receiving NNRTI-based regimens. Substantial geographic heterogeneity was observed, with viral load uptake varying across regions and districts. Several districts characterized by mobile populations and seasonal migration patterns demonstrated comparatively lower monitoring coverage.

**Conclusions:** Although viral load monitoring coverage and suppression levels among those tested were relatively high, important gaps remain in testing uptake and geographic equity. Adolescents, young adults, and populations in certain districts experience lower monitoring coverage and treatment outcomes. Strengthening viral load monitoring systems, expanding youth-responsive HIV services, and addressing geographic disparities will be important for sustaining progress toward national and global HIV treatment targets.

## Introduction

Timely viral load monitoring is crucial for achieving the UNAIDS 95-95-95 targets and sustaining long-term outcomes of antiretroviral therapy. Routine viral load testing is the preferred approach for assessing treatment response, identifying virological failure early, and guiding adherence interventions or regimen switches. Despite the expansion of access to viral load testing in many sub-Saharan African countries, significant gaps persist due to logistical, programmatic, and patient-level challenges that limit progress toward epidemic control [1,2].

Tanzania adopted routine viral load monitoring in line with the World Health Organization (WHO) recommendations under the leadership of the National AIDS and Sexually Transmitted Infections Control Programme (NASHCoP). The expansion of antiretroviral (ART) coverage and viral load laboratory capacity has strengthened national monitoring systems. However, many people living with HIV continue to miss viral load testing at the recommended six- and twelve-month intervals after ART initiation, which undermines early detection of treatment failure and reduces the overall effectiveness of treatment programmes [3].

Evidence from Tanzania has shown the importance of timely monitoring. A landmark study demonstrated that pharmacy refill adherence strongly predicted virological failure among adults in Dar es Salaam [4], highlighting the need for high-quality monitoring across the care continuum. Regional studies consistently describe disparities in viral load, uptake, and suppression by age, gender, regimen category, and clinical status. Multi-country analyses from East Africa report lower viral load uptake and poorer suppression among adolescents, young adults, and men [5,6]. Studies from Uganda have further indicated persistently low viral load monitoring coverage among adolescents [7]. Longitudinal data from Southern Africa indicate improvements in viral load uptake over time; however, persistent inequities persist among younger age groups [8]. Psychosocial vulnerabilities have also been associated with virological non-suppression in adolescent populations [9].

The transition to dolutegravir-based regimens has contributed to improved programme outcomes, driven by their favorable safety profile and high genetic barrier to resistance. Clinical trial data and implementation reports across African programmes show enhanced viral suppression following dolutegravir rollout [10,11]. Differentiated service delivery models have complemented these improvements by enhancing continuity and reducing the clinical burden among stable patients [12].

Despite these advances, comprehensive national analyses describing viral load testing uptake, viral suppression, and geographic variation across Tanzania remain limited. Most existing reports are regional or facility-based and do not fully capture nationwide programme performance. Using routinely collected national programme data from the NASHCoP CTC-2 database, this study examined viral load testing uptake, viral suppression, and associated demographic, treatment, and geographic predictors among people receiving antiretroviral therapy in Tanzania between 2017 and 2021. The findings are intended to guide targeted interventions to strengthen routine viral load monitoring and improve equitable programme performance. essential.

## Methods

### Study design

This study employed a retrospective observational design using routinely collected HIV programme data to evaluate viral load monitoring and treatment outcomes among people living with HIV receiving antiretroviral therapy in Tanzania. The analysis focused on patterns of viral load testing uptake, viral suppression, and associated demographic, treatment, and geographic predictors during 2017-2021.

### Data source

Data were obtained from the Care and Treatment Clinic electronic database (CTC-2), maintained by the National AIDS and Sexually Transmitted Infections Control Programme (NASHCoP) of the Ministry of Health in Tanzania. The CTC-2 system captures routine clinical, demographic, treatment, and laboratory information for individuals receiving HIV care and treatment services across mainland Tanzania.

The dataset used for this analysis consisted of a national extract of anonymized patient records covering the period from 1^st^ January 2017 to 31^st^ December 2021. The dataset included demographic information, ART regimen history, clinic visit dates, viral load test records, pharmacy refill adherence indicators, and administrative location variables, including district and region of care.

### Study setting

The analysis covered all 26 mainland Tanzania regions and their corresponding administrative districts. In Tanzania, HIV care services are delivered through care and treatment clinics located in dispensaries, health centers, district hospitals, regional referral hospitals, and specialized facilities.

Because of the large number of facilities included in the national dataset, analyses were conducted at the regional and district levels rather than at the individual facility level. This approach allowed assessment of geographic variation in viral load uptake and treatment outcomes while maintaining analytical feasibility.

### Study population

The study population consisted of 70,000 individuals receiving antiretroviral therapy who had valid demographic information and treatment records within the national programme dataset during the study period.

Individuals were included if they had available data on age, gender, ART regimen, clinic visit records, and geographic identifiers (district and region). Records with missing key demographic variables or incomplete geographic information were excluded during data cleaning.

The analysis included children, adolescents, and adults who received ART during the observation window. Because the dataset was extracted within a defined time window rather than constructed as an incident-treatment cohort, the first appearance of an individual in the dataset represents the first recorded observation within the study period, rather than necessarily the first lifetime ART initiation date.

### Variables and definitions

Demographic variables included age, gender, and geographic location of care (district or region). Age groups were categorized into 0-10 years, 11-18 years, 19-28 years, 29-69 years, and ≥70 years based on the cohort distribution.

Treatment variables included information on the ART regimen at the first and last observed visits during the study period. The regimens were classified into major regimen classes based on the anchor drug component, including dolutegravir**-**based regimens (DTG), Non-nucleoside reverse transcriptase inhibitor regimens (NNRTI), protease inhibitor regimens (PI) and other regimens.

Pharmacy refill adherence was calculated using pharmacy dispensing records and expressed as a percentage, representing the proportion of days covered by dispensed medication over the observation period.

The primary outcome was viral load testing uptake, which was defined as the presence of at least one recorded viral load result in the dataset during the study period.

The secondary outcome was viral load suppression, defined as a viral load measurement <1,000 copies/mL among individuals with at least 1 documented viral load test.

### Data processing

Data cleaning and preparation were conducted in the R statistical environment. Records were inspected for missing values, implausible entries, and duplicate observations before analysis.

Categorical variables were standardized, and ART regimens were grouped into regimen classes based on their anchor drug component. Geographic identifiers were harmonized to ensure consistency with national administrative districts and regional boundaries used for spatial analysis.

District-level and regional-level summary datasets were generated to facilitate descriptive analyses and geographic visualization.

### Statistical analysis

Descriptive statistics were used to summarize the baseline demographic and treatment characteristics of the study population. Continuous variables are summarized using means and standard deviations, whereas categorical variables are summarized using frequencies and percentages.

The uptake of viral load testing and viral suppression was calculated overall and stratified by demographic characteristics, regimen category, and geographic location.

Geographic variations in viral load uptake and adherence were visualized using choropleth maps generated from national administrative boundary shapefiles. Regional and district-level summary tables were generated to accompany these visualizations.

Multivariable logistic regression models were used to identify the predictors of viral load testing uptake and viral load suppression among individuals with available viral load results.

The covariates included age group, gender, regimen class, pharmacy refill adherence category, calendar year of the first observation in the dataset, and geographic region.

Adjusted odds ratios (aOR) with 95% confidence intervals are reported for all predictors.

### Ethical considerations

Ethical approval for this study was obtained from the Muhimbili University of Health and Allied Sciences Research Ethics Committee (MUHAS-IRB) under reference number DA.282/298/01.C/MUHAS-REC-10-2020-379.

Permission to access and analyze anonymized programme data was granted by **the** National AIDS and Sexually Transmitted Infections Control Programme (NASHCoP).

All data were de-identified prior to analysis, and no personal identifiers were retained in the analytical dataset.

## Results

### Study population

We included 70, 000 people living with HIV who were receiving antiretroviral therapy in the analysis. The mean age at initiation of ART was 37.0 years (SD 13.4) and 39.2 years (SD 13.7) at the end of the observation period. The mean follow-up duration was 939.6 days (SD 678.0), with a mean of 19.3 clinic visits (SD 13.9) and 0.9 therapy changes (SD 0.9). Women comprised 45,561 (65.1%) of the participants. Adults aged 29-69 years represented the largest age group (51,783; 74.0%). The largest regional contributions were from Dar es Salaam, Mwanza, and Mbeya (Table 1).

**Table 1:**
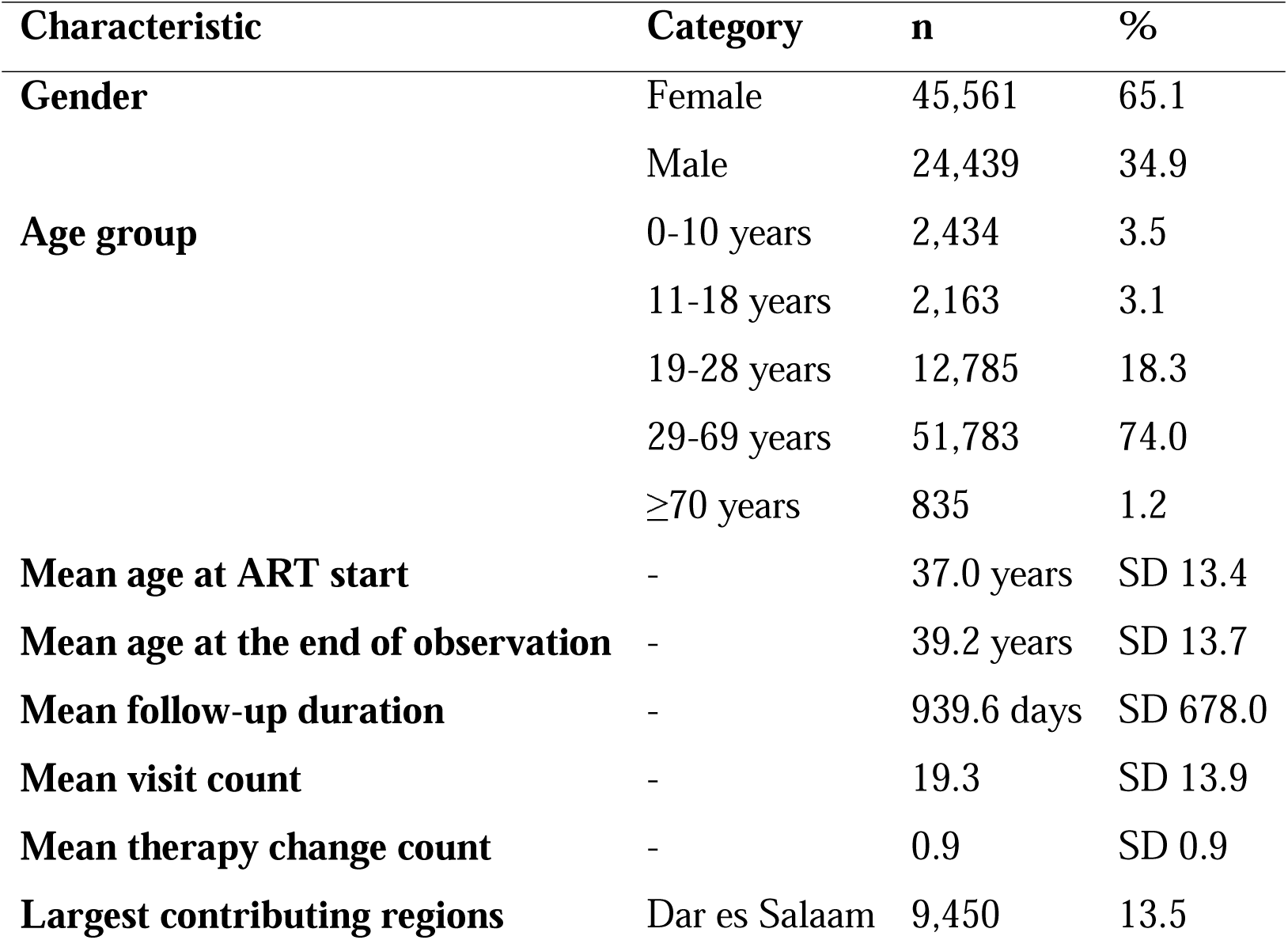
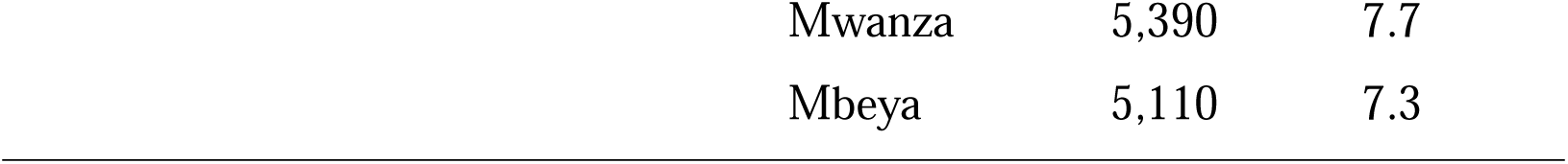
Baseline demographic and clinical characteristics of people living with HIV receiving antiretroviral therapy in Tanzania, 2017-2021 (n = 70,000)

| Characteristic | Category | n | % |
| --- | --- | --- | --- |
| **Gender** | Female | 45,561 | 65.1 |
|  | Male | 24,439 | 34.9 |
| **Age group** | 0-10 years | 2,434 | 3.5 |
|  | 11-18 years | 2,163 | 3.1 |
|  | 19-28 years | 12,785 | 18.3 |
|  | 29-69 years | 51,783 | 74.0 |
|  | ≥70 years | 835 | 1.2 |
| **Mean age at ART start** | - | 37.0 years | SD 13.4 |
| **Mean age at the end of observation** | - | 39.2 years | SD 13.7 |
| **Mean follow-up duration** | - | 939.6 days | SD 678.0 |
| **Mean visit count** | - | 19.3 | SD 13.9 |
| **Mean therapy change count** | - | 0.9 | SD 0.9 |
| **Largest contributing regions** | Dar es Salaam | 9,450 | 13.5 |

|  |  |  |
| --- | --- | --- |
| Mwanza | 5,390 | 7.7 |
| Mbeya | 5,110 | 7.3 |

### ART regimen patterns

For the first recorded regimen, NNRTI-based therapy predominated (52,227; 74.6%), followed by dolutegravir-based regimens (15,877; 22.7%), protease inhibitor regimens (1,420; 2.0%), and other regimens (64; 0.1%), with 0.7% unknown. The most common initial regimen was TDF-3TC-EFV (41,043; 58.6%), followed by TDF-3TC-DTG (15,726; 22.5%) (Table 2).

**Table 2:** Distribution of antiretroviral therapy regimens at the first and last observed visits among people living with HIV receiving treatment in Tanzania, 2017-2021.

Regimen class distribution
| Regimen class | First regimen n (%) | Last regimen n (%) |
| --- | --- | --- |
| DTG-based | 15,890 (22.7) | 51,450 (73.5) |
| NNRTI-based | 52,220 (74.6) | 15,960 (22.8) |
| PI-based | 1,400 (2.0) | 2,030 (2.9) |
| Other | 70 (0.1) | 140 (0.2) |
| Unknown | 420 (0.7) | 420 (0.7) |

Most common individual regimens
| Regimen | First regimen % | Last regimen % |
| --- | --- | --- |
| TDF + 3TC + EFV | 58.6 | 19.8 |
| TDF + 3TC + DTG | 22.5 | 72.3 |
| AZT + 3TC + NVP | 8.5 | <1 |

### Regimen class distribution

By the last recorded regimen, DTG-based therapy had become dominant (51,456; 73.5%), whereas NNRTI-based regimens accounted for 15,976 (22.8%). The most common final regimen was TDF-3TC-DTG (50,611; 72.3%), followed by TDF-3TC-EFV (13,860; 19.8%) (Table 2).

### Viral load uptake

Overall, 49,820 (71.2%) individuals had at least one documented viral load measurement during the study period, whereas 20,180 (28.8%) had none. Pharmacy refill adherence data were available for 64,803 (92.6%) individuals, with a mean adherence of 85.0%. Summary indicators for viral load testing and monitoring are presented in Table 3.

**Table 3:** Viral load testing uptake and availability of monitoring indicators among people living with HIV receiving antiretroviral therapy in Tanzania, 2017-2021.

| **Indicator** | **n** | **%** |
| --- | --- | --- |
| Viral load result available | 49,820 | 71.2 |
| No viral load result recorded | 20,180 | 28.8 |
| Adherence data available | 64,803 | 92.6 |
| No adherence data | 5,197 | 7.4 |
| **Mean adherence** | - | 85.0% |

Viral load uptake differed across demographic groups. Uptake was 75.3% among adolescents aged 11-18 years, 73.7% among children aged 0-10 years, 73.6% among adults aged 29-69 years, 67.2% among elderly individuals aged ≥70 years, and 60.5% among young adults aged 19-28 years. Uptake was slightly higher among females (72.2%) than among males (69.3%).

When uptake was stratified by first-observed year within the dataset, it was highest among individuals first observed in 2017 (82.6%), followed by 2020 (68.6%), 2019 (65.0%), and 2018 (64.7%), while those first observed in 2021 had lower uptake (34.4%). Uptake also differed by regimen class, with higher uptake among individuals receiving DTG-based (83.7%) and PI-based (82.0 %) regimens compared with NNRTI-based regimens (31.2%) (Table 3).

### Viral suppression

Among the 49,820 individuals with documented viral load results, 44,187 achieved viral suppression, while 5,633 did not, yielding an overall suppression rate of 88.7% (Table 4).

**Table 4:** Viral load suppression among individuals with documented viral load results, stratified by demographic and treatment characteristics (n = 49,820)

| Category | Subgroup | Suppressed n (%) | Not suppressed n (%) |
| --- | --- | --- | --- |
| **Overall** | - | 44,187 (88.7) | 5,633 (11.3) |
| **Sex** | Female | 29,383 (89.4) | 3,501 (10.6) |
|  | Male | 14,804 (87.4) | 2,132 (12.6) |
| **Age group at ART start** | Children (0-10) | 1,226 (68.3) | 568 (31.7) |
|  | Adolescents (11-18) | 1,124 (69.0) | 505 (31.0) |
|  | Young adults (19-28) | 6,996 (90.4) | 741 (9.6) |
|  | Adults (29-69) | 34,339 (90.1) | 3,760 (9.9) |
| | Elderly ($\geq 70$) | 502 (89.5) | 59 (10.5) |
| **Last regimen class** | DTG-based | 39,212 (91.0) | 3,874 (9.0) |
|  | NNRTI-based | 3,978 (79.9) | 1,002 (20.1) |
|  | PI-based | 940 (57.4) | 697 (42.6) |
|  | Other | 53 (48.6) | 56 (51.4) |
|  | Unknown | 4 (50.0) | 4 (50.0) |

Suppression levels differed across age groups. Suppression was 69.0% among adolescents aged 11-18 years and 68.3% among children aged 0-10 years, compared with 90.1% among adults aged 29-69 years, 90.4% among young adults aged 19-28 years, and 89.5% among elderly individuals aged ≥70 years. Suppression was slightly lower among males (87.4%) than among females (89.4%). By regimen class, suppression was highest among those using DTG-based regimens (91.0%), followed by NNRTI-based regimens (79.9%), PI-based regimens (57.4%), and other regimens (48.6%) (Table 4).

### Adherence availability by viral load status

Adherence information was available for 15,287 (75.8%) individuals without documented viral load results and for 49,516 (99.4%) of those with viral load measurements. The availability of adherence information by viral load status (Table 5).

**Table 5:** Availability of pharmacy refill adherence information according to viral load testing status among people receiving antiretroviral therapy in Tanzania.

| Viral load status | Adherence available n | Adherence missing n |
| --- | --- | --- |

|  | (%) | (%) |
| --- | --- | --- |
| Viral load documented (n=49,820) | 49,516 (99.4) | 304 (0.6) |
| No viral load documented (n=20,180) | 15,287 (75.8) | 4,893 (24.2) |

### Geographic variation in viral load uptake and suppression

Regional variations in viral load uptake were observed across the country. Uptake ranged from 62.4% in Manyara to 76.6% in Mbeya, with relatively high uptake also observed in Iringa (76.3%), Mara (76.3%), Mwanza (75.3%), and Songwe (74.5%). Lower uptake was observed in Katavi (63.7%), Geita (66.0%), Singida (65.7%), Arusha (66.2%), and Morogoro (66.8%). Regional suppression among those tested ranged from 78.0% in Ruvuma to 93.7% in Geita. In contrast, the mean adherence ranged from 78.5% in Rukwa to 89.6% in Iringa (Table 6), consistent with the di spatial patterns (Figure 1).

**Figure 1:**
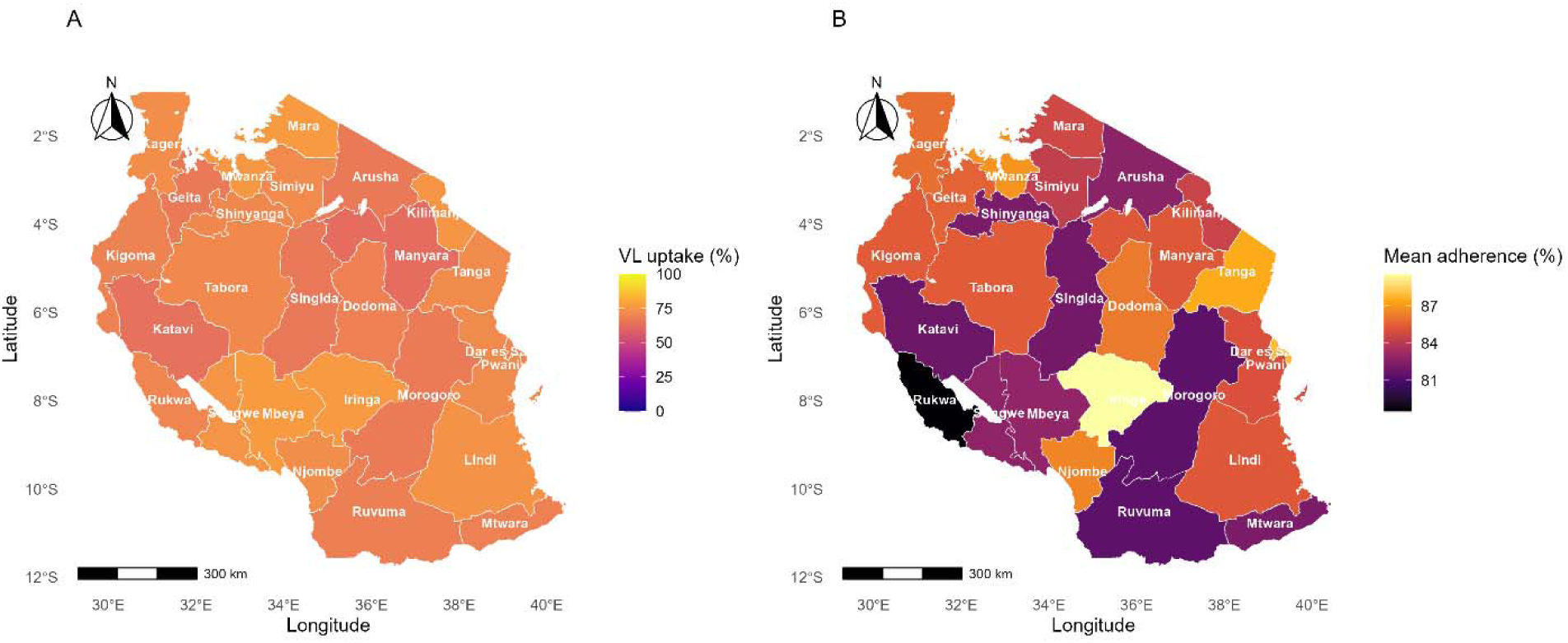
Regional variations in viral load uptake and mean pharmacy refill adherence among people living with HIV receiving antiretroviral therapy in Tanzania, 2017-2021. Panel A: Viral load testing uptake by region. Panel B: Mean pharmacy refill adherence by region.

**Table 6:** Viral load suppression among individuals with documented viral load results, stratified by demographic and treatment characteristics (n = 49,820)

| Region | VL uptake % | VL suppression % | Mean adherence % |
| --- | --- | --- | --- |
| Arusha | 66.2 | 82.6 | 82.7 |
| Dar es Salaam | 69.6 | 86.1 | 87.9 |
| Dodoma | 68.2 | 85.9 | 86.2 |
| Geita | 66.0 | 93.7 | 85.6 |
| Iringa | 76.3 | 88.1 | 89.6 |
| Kagera | 72.6 | 92.2 | 85.9 |
| Katavi | 63.7 | 91.7 | 81.8 |
| Kigoma | 68.7 | 90.4 | 85.4 |
| Kilimanjaro | 74.7 | 83.8 | 84.5 |
| Lindi | 73.7 | 81.8 | 85.3 |
| Manyara | 62.4 | 85.1 | 85.3 |
| Mara | 76.3 | 89.5 | 84.7 |
| Mbeya | 76.6 | 89.2 | 82.9 |
| Morogoro | 66.8 | 86.3 | 81.4 |
| Mtwara | 68.6 | 84.8 | 82.2 |
| Mwanza | 75.3 | 92.9 | 86.9 |
| Njombe | 72.4 | 90.0 | 86.5 |
| Pwani | 71.2 | 87.9 | 85.1 |
| Rukwa | 69.8 | 84.3 | 78.5 |
| Ruvuma | 67.8 | 78.0 | 81.4 |
| Shinyanga | 70.2 | 93.0 | 82.2 |
| Simiyu | 71.3 | 91.9 | 84.3 |
| Singida | 65.7 | 89.2 | 82.0 |
| Songwe | 74.5 | 92.8 | 82.8 |
| Tabora | 71.1 | 92.0 | 85.4 |
| Tanga | 71.4 | 86.4 | 87.5 |

### District-level variation

District-level summaries showed heterogeneity within regions. In the Arusha Region, viral load uptake ranged from 41.7% in Ngorongoro and 48.3% in Longido to 75.9% in Arumeru. In the Manyara Region, uptake was 49.0% in Simanjiro and 56.4% in Kiteto. In the Singida Region, Ikungi recorded a 48.2% uptake. In the Ruvuma Region, uptake was 53.1% in Namtumbo and 57.8% in Tunduru, while in the Tanga Region, uptake was 60.1% in Kilindi. District-level spatial patterns (Figure 2).

**Figure 2:**
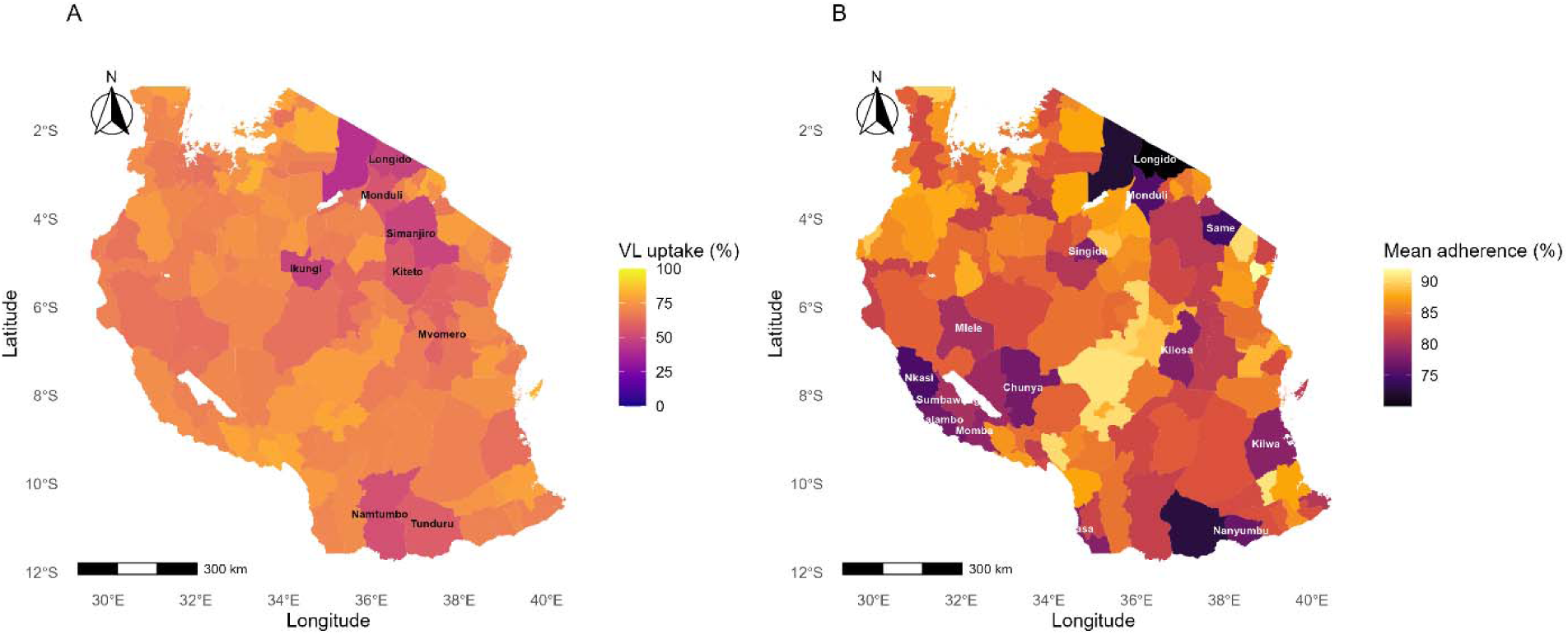
District-level variation in viral load uptake and mean pharmacy refill adherence among people living with HIV receiving antiretroviral therapy in Tanzania, 2017-2021.

### Panel A: Viral load testing uptake by district

Panel A: Viral load testing uptake by district. Panel B: Mean pharmacy refill adherence by district. Districts with viral load uptake below 60% are labeled.

### Temporal variation by region

Across regions, viral load uptake was the highest among individuals first observed in 2017 and lower among those first observed in 2021. For example, the uptake declined from 84.6% to 38.4% in Mbeya, 84.3% to 33.6% in Mwanza, and 84.0% to 13.0% in Ruvuma between these observation years (Supplementary Table 1).

### Predictors of viral load uptake

In the multivariate logistic regression analysis, viral load uptake was associated with age, gender, first-observed year, adherence status, regimen class, and region (Table 7). Compared with adolescents aged 11-18 years, the odds of viral load uptake were higher among adults aged 29-69 years (aOR 3.53, 95% CI 3.14-3.97), individuals aged ≥70 years (aOR 3.35, 95% CI 2.50-4.57), young adults aged 19-28 years (aOR 3.10, 95% CI 2.70-3.56), and children aged 0-10 years (aOR 1.35, 95% CI 1.15-1.58).

**Table 7:** Multivariable logistic regression analysis of factors associated with viral load uptake among people living with HIV receiving antiretroviral therapy in Tanzania, 2017-2021.

| Variable | Category | aOR | 95% CI | p-value |
| --- | --- | --- | --- | --- |
| **Age group at ART start** | Adolescents (11-18) | Reference | - | - |
|  | Children (0-10) | 1.35 | 1.01-1.50 | 0.043 |
|  | Young adults (19-28) | 3.10 | 2.70-3.56 | <0.001 |
|  | Adults (29-69) | 3.53 | 3.14-3.97 | <0.001 |
| | Elderly ($\geq 70$) | 3.35 | 2.50-4.57 | <0.001 |
| **Sex** | Female | Reference | - | - |
|  | Male | 0.83 | 0.78-0.88 | <0.001 |
| **Adherence group** | <85% | Reference | - | - |
| | $\geq 85\%$ | 1.72 | 1.62-1.83 | <0.001 |
| **Regimen class** | DTG-based | Reference | - | - |
|  | NNRTI-based | 0.51 | 0.47-0.56 | <0.001 |
|  | PI-based | 0.20 | 0.18-0.23 | <0.001 |
|  | Other | 0.12 | 0.08-0.18 | <0.001 |
| **First observed year** | 2017 | Reference | - | - |
|  | 2018 | 0.42 | 0.39-0.45 | <0.001 |
|  | 2019 | 0.16 | 0.14-0.17 | <0.001 |
|  | 2020 | 0.07 | 0.06-0.08 | <0.001 |
|  | 2021 | 0.01 | 0.01-0.02 | <0.001 |

Male gender was associated with lower odds of viral load uptake compared with females (aOR 0.83, 95% CI 0.78-0.88). Individuals with adherence ≥85% had higher odds of viral load uptake than those with adherence <85% (aOR 1.72, 95% CI 1.62-1.83).

Compared with DTG-based regimens, viral load uptake was lower among individuals receiving NNRTI-based regimens (aOR 0.51, 95% CI 0.47-0.56), PI-based regimens (aOR 0.20, 95% CI 0.18-0.23), and other regimens (aOR 0.12, 95% CI 0.08-0.18).

### Predictors of viral suppression

Among individuals with documented viral load results, viral suppression was associated with demographic, treatment, and programmatic characteristics (Table 8). Male gender was associated with lower odds of suppression compared with females (aOR 0.84, 95% CI 0.80-0.88).

**Table 8:** Multivariable logistic regression analysis of factors associated with viral suppression among individuals with documented viral load results in Tanzania.

| Variable | Category | aOR | 95% CI | p-value |
| --- | --- | --- | --- | --- |
| Age group at ART start | Adolescents (11-18) | Reference | - | - |
|  | Children (0-10) | 1.23 | 1.01-1.50 | 0.0003 |
|  | Young adults (19-28) | 0.72 | 0.62-0.83 | <0.001 |
|  | Adults (29-69) | 3.53 | 3.14-3.97 | <0.001 |
| | Elderly ($\geq 70$) | 3.35 | 2.50-4.57 | <0.001 |
| Sex | Female | Reference | - | - |
|  | Male | 0.84 | 0.80-0.88 | <0.001 |
| Adherence information | No adherence data | Reference | - | - |
|  | Adherence documented | 19.18 | 16.81-21.97 | <0.001 |
| Regimen class | DTG-based | Reference | - | - |
|  | NNRTI-based | 0.023 | 0.021-0.024 | <0.001 |
|  | PI-based | 0.36 | 0.31-0.43 | <0.001 |

Young adults aged 19-28 years had lower odds of suppression (aOR 0.72, 95% CI 0.62-0.83) than adolescents aged 11-18 years, whereas children aged 0-10 years had slightly higher odds (aOR 1.23, 95% CI 1.01-1.50).

Compared with DTG-based regimens, viral suppression was lower among individuals receiving NNRTI-based regimens (aOR 0.023, 95% CI 0.021-0.024) and PI-based regimens (aOR 0.36, 95% CI 0.31-0.43).

Individuals with documented adherence information had higher odds of viral suppression than those without adherence data (aOR 19.18, 95% CI 16.81-21.97).

## Discussion

This national analysis of routine HIV programme data provides an overview of viral load monitoring and treatment outcomes among people receiving antiretroviral therapy in Tanzania between 2017 and 2021. Four main findings emerge. Four main findings emerged: First, although viral load testing coverage was substantial, nearly one-third of patients lacked documented viral load results during the observation period, indicating persistent gaps in routine monitoring. Such gaps in the viral load monitoring cascade are not unique to Tanzania but represent a systemic challenge across sub-Saharan Africa [13], often complicated by varying definitions and criteria used to measure progress within the care cascade [14] Second, important demographic differences were observed: the lowest viral load uptake was among young adults, while viral suppression was lower among children and adolescents than among older adults. Men also performed less well than women across several indicators. Third, patients receiving dolutegravir-based regimens had higher viral suppression than those on older regimen classes [15]. Fourth, marked geographic heterogeneity was observed across regions and districts, indicating that local health-system conditions and continuity-of-care challenges may influence programme performance.

Substantial geographic heterogeneity in viral load uptake was observed across regions and districts. Similar spatial variation in HIV programme indicators has been reported in multi-country analyses of population viral load and treatment coverage across Africa [16]. These patterns likely reflect differences in health system capacity, access barriers, laboratory infrastructure, and population mobility. Recent evidence suggests that even with improved drug regimens, biological and social predictors-including distance to facility and regional socioeconomic status-continue to drive inequities in suppression across the region [17]. Spatial analyses conducted in other African settings have also shown that HIV programme performance often clusters geographically, reflecting underlying socioeconomic and structural determinants [18,19].

An important observation from the spatial analysis was that lower viral load testing uptake appeared more clearly clustered within mobility-prone districts than pharmacy refill adherence. Several northern pastoralist and southern border districts had relatively low viral load monitoring coverage, whereas the adherence map showed a weaker, less consistent spatial signal. This pattern suggests that, in some settings, the continuity of viral load monitoring may be more vulnerable to population mobility, silent transfer, challenges in specimen transport, or fragmented documentation than medication pickup behavior alone. These observations are consistent with findings from analyses of routine HIV programme data, which show that continuity of care indicators can be influenced by population mobility and health system documentation patterns in large national datasets [20,21]. Together, these findings suggest that some routine monitoring gaps may partly reflect continuity-of-care and recording challenges rather than poor treatment adherence alone. Spatial clustering of programme indicators has also been observed in geospatial analyses of ART consumption across Tanzania [22].

Age-related disparities were also evident, although they differed by outcome. Viral load uptake was lowest among young adults, whereas viral suppression was lower among children and adolescents than among older adults. This suggests that different mechanisms may underlie these patterns. Among young adults, lower uptake may reflect weaker engagement with routine monitoring, transitions in care, and competing social or economic demands. Similar age-specific inequities and correlates of poor suppression have recently been documented among adolescents and young adults in urban centers such as Dar es Salaam, underscoring the need for youth-friendly monitoring strategies [23]. Among adolescents and children, poorer suppression may additionally reflect adherence challenges, caregiver dependence, disclosure difficulties, psychosocial vulnerability, and transitions between pediatric and adult HIV services. These findings are consistent with regional evidence showing persistent challenges with treatment and viral suppression among younger populations living with HIV in sub-Saharan Africa [6,7,9,24].

Patients receiving dolutegravir-based regimens demonstrated higher viral suppression compared with those receiving older NNRTI-based regimens. These findings align with clinical trial evidence and programme-level analyses demonstrating superior virological outcomes with dolutegravir-based therapy [10,11,15,25]. The introduction of dolutegravir across African HIV programmes has been associated with improved viral suppression and greater regimen durability. This is consistent with recent findings from Mozambique, where the pediatrics DTG rollout led to high rates of virological suppression, though challenges remain in specific age subsets [26]. However, persistent disparities among adolescents and younger adults suggest that pharmacological improvements alone are insufficient to close remaining gaps in treatment outcomes.

This analysis complements a growing body of work examining HIV programme performance in Tanzania using routinely collected national datasets. Previous studies have investigated predictors of loss to follow-up, geographic variation in retention patterns, and the role of pharmacy refill adherence in predicting treatment outcomes [21,27,28]. Additional analyses using national programme data have also demonstrated substantial geographic variation in loss to follow-up and highlighted the potential utility of machine-learning approaches for predicting virological outcomes using routinely collected clinical indicators [29]. Taken together, these studies illustrate how routine programme data can provide important insights into both clinical outcomes and health-system performance within national HIV programmes.

Future work integrating viral load monitoring with pharmacy refill data, appointment-keeping behavior, and other electronic health records could further strengthen programme monitoring systems. Addressing the leaks in the viral load monitoring cascade is essential, as systematic reviews confirm that the transition from clinical eligibility for a test to actually receiving a result remains the weakest link in many SSA programmes [13]. Machine-learning approaches applied to routine programme datasets may help identify patients at risk of virological failure or disengagement from care, enabling earlier and more targeted interventions [29].

### Strengths and limitations

A key strength of this study is the use of a large national programme dataset covering all regions and districts of mainland Tanzania over multiple years. This enabled a broad assessment of viral load uptake, viral suppression, and spatial heterogeneity in programme performance. The analysis extends previous regional studies by incorporating district-level summaries and geographic visualization, thereby identifying areas where programme gaps may be concentrated.

Several limitations should also be considered. First, as with all routine programme datasets, data completeness and quality may vary across facilities and districts, and missing viral load results may have influenced estimates of monitoring coverage and suppression. Second, because the dataset represents a programme extract within a defined observation window, the first recorded visit does not necessarily correspond to the first lifetime ART initiation. Third, behavioral and social determinants, such as stigma, mental health, disclosure, and socioeconomic status, were not available. Fourth, although geographic patterns were examined, the analysis did not include direct measures of population mobility, facility-level service characteristics, laboratory turnaround time, or specimen transport performance. As a result, spatial patterns should be interpreted cautiously as indicators of possible programme and continuity-of-care differences rather than direct evidence of causation.

### Implications for policy and practice

These findings have several implications for Tanzania’s HIV programme. First, continued efforts are required to improve viral load testing coverage, particularly among adolescents, young adults, and men. Strengthening specimen transport systems, improving turnaround time for laboratory results, and integrating viral load monitoring into differentiated service delivery models could help close these gaps.

Second, the persistent suppression gap among adolescents highlights the need for adolescent-friendly HIV services that address adherence challenges, psychosocial support, and transitions between pediatric and adult care. Targeted interventions addressing stigma, school-related barriers, and mental health support may also improve outcomes in younger populations.

Third, the transition to dolutegravir-based regimens should continue to be prioritized while maintaining careful monitoring for treatment adherence and potential drug resistance. Programme managers can also use district-level maps and summary tables generated in this analysis to identify underperforming areas and prioritize technical support, mentorship, and resource allocation.

Finally, integrating pharmacy refill adherence data and other routinely collected programme indicators into predictive models may help identify individuals at risk of treatment failure or missed viral load monitoring. Such approaches may enable proactive interventions, particularly in resource-limited settings where universal viral load testing coverage may remain challenging.

In districts with lower viral load monitoring but moderate adherence, programme managers may need to examine continuity-of-care pathways, silent transfer patterns, specimen transport systems, and local recording practices in addition to adherence support.

## Conclusion

This national analysis of routine HIV programme data shows that viral load monitoring coverage in Tanzania was substantial during 2017-2021 and that viral suppression among those tested was high. However, important gaps remain in monitoring coverage and geographic equity. Young adults had the lowest viral load uptake, while children and adolescents showed poorer viral suppression than older adults. Marked district-level heterogeneity suggests that health system conditions, continuity-of-care challenges, and population mobility may influence routine monitoring patterns. Strengthening viral load monitoring systems, expanding youth-responsive HIV services, and addressing geographic disparities will be important for achieving more equitable treatment outcomes and sustaining progress toward national and global HIV targets.

## Declaration

## Acknowledgements

We thank the National AIDS and Sexually Transmitted Infections Control Programme (NASHCoP) of the Ministry of Health, Tanzania, for granting access to anonymized CTC-2 program data used in this analysis. We also acknowledge the regional and district health management teams, as well as frontline healthcare workers, who contributed to the routine collection and reporting of HIV data. We are grateful to the data officers who maintain the CTC-2 system and ensure the integrity of the national programme records.

## Data availability statement

The data used in this study are routinely collected HIV programme records owned by the National AIDS and Sexually Transmitted Infections Control Programme (NASHCoP), Ministry of Health, Tanzania. These data are not publicly available because of confidentiality restrictions. De-identified datasets may be made available by NASHCoP upon reasonable request and are subject to national data-sharing policies.

## Ethics approval

Ethical approval for this study was obtained from the Muhimbili University of Health and Allied Sciences Research Ethics Committee (MUHAS-IRB), Ref: DA.282/298/01.C/MUHAS-REC-10-2020-379. Permission to access and analyze anonymized CTC-2 programme data was granted by the National AIDS and Sexually Transmitted Infections Control Programme (NASHCoP). All data were de-identified prior to analysis, and no personal identifiers were retained.

## Supplementary Table

**Supplementary Table 1:** Temporal variation in viral load uptake by region and first observed year in the dataset window, Tanzania HIV programme data (2017-2021)

| Region | 2017 Uptake % | 2018 Uptake % | 2019 Uptake % | 2020 Uptake % | 2021 Uptake % |
| --- | --- | --- | --- | --- | --- |
| Arusha | 82.7 | 64.9 | 65.8 | 69.3 | 36.1 |
| Dar es Salaam | 83.9 | 65.7 | 66.4 | 70.1 | 35.2 |
| Dodoma | 83.4 | 65.2 | 66.0 | 70.5 | 35.8 |
| Geita | 83.1 | 64.4 | 65.1 | 69.6 | 34.2 |
| Iringa | 84.4 | 66.3 | 67.1 | 71.4 | 38.7 |
| Katavi | 82.2 | 63.8 | 64.9 | 68.3 | 33.5 |
| Kigoma | 82.5 | 64.2 | 65.0 | 68.7 | 33.9 |
| Kilimanjaro | 83.6 | 65.5 | 66.6 | 70.3 | 35.6 |
| Lindi | 82.4 | 63.7 | 64.8 | 68.1 | 33.0 |
| Manyara | 82.8 | 64.1 | 65.3 | 69.0 | 34.6 |
| Mara | 84.1 | 66.2 | 67.0 | 71.0 | 37.9 |
| Mbeya | 84.6 | 66.8 | 67.4 | 71.8 | 38.4 |
| Morogoro | 83.0 | 65.1 | 66.0 | 69.7 | 34.9 |
| Mtwara | 82.6 | 64.0 | 64.9 | 68.5 | 33.7 |
| Mwanza | 84.3 | 66.4 | 67.2 | 71.2 | 33.6 |
| Njombe | 84.2 | 66.6 | 67.3 | 71.5 | 37.8 |
| Pwani | 83.3 | 65.3 | 66.1 | 70.0 | 35.0 |
| Rukwa | 82.1 | 63.5 | 64.4 | 68.0 | 32.8 |
| Ruvuma | 84.0 | 65.8 | 66.5 | 70.6 | 13.0 |
| Shinyanga | 83.5 | 65.0 | 65.8 | 69.4 | 34.1 |
| Singida | 82.9 | 64.6 | 65.4 | 69.1 | 34.3 |
| Songwe | 84.1 | 66.5 | 67.2 | 71.1 | 37.6 |
| Tabora | 82.7 | 64.3 | 65.1 | 68.8 | 33.8 |
| Tanga | 83.2 | 65.0 | 65.9 | 69.6 | 34.7 |

## References

1. UNAIDS. The path that ends AIDS: UNAIDS Global AIDS Update 2023. 2023 [cited 10 Mar 2026]. Available: https://www.unaids.org/en/resources/documents/2023/global-aids-update-2023

2. United Republic of Tanzania. Tanzania HIV Impact Survey 2022-2023. 2024 [cited 18 Dec 2025]. Available: https://www.nbs.go.tz/nbs/takwimu/THIS2022-2023/THIS2022-2023_Summary_Sheet.pdf

3. World Health Organization. Consolidated guidelines on HIV prevention, testing, treatment, service delivery and monitoring: recommendations for a public health approach. 2021 [cited 10 Mar 2021]. Available: https://www.who.int/publications/i/item/9789240031593

4. Sangeda RZ, Mosha F, Prosperi M, Aboud S, Vercauteren J, Camacho RJ, et al. Pharmacy refill adherence outperforms self-reported methods in predicting HIV therapy outcome in resource-limited settings. BMC Public Health. 2014;14: 1035. doi:10.1186/1471-2458-14-1035

5. Adhiambo HF, Lewis-Kulzer J, Nyagesoa E, Gimbel S, Akama E, Mangale D, et al. Examining and classifying reasons for missing viral loads among adults living with HIV: An extended outcome investigation and ascertainment approach in Western Kenya. Goyal, V, editor. PLOS Global Public Health. 2025;5: e0004038. doi:10.1371/journal.pgph.0004038

6. Quaker AS, Shirima LJ, Msuya SE. Trend and factors associated with non-suppression of viral load among adolescents on ART in Tanzania: 2018–2021. Frontiers in Reproductive Health. 2024;6. doi:10.3389/frph.2024.1309740

7. Nasuuna E, Kigozi J, Babirye L, Muganzi A, Sewankambo NK, Nakanjako D. Low HIV viral suppression rates following the intensive adherence counseling (IAC) program for children and adolescents with viral failure in public health facilities in Uganda. BMC Public Health. 2018;18: 1048. doi:10.1186/s12889-018-5964-x

8. Otto M, Okango E, Mee P, Dobra A, Tram KH, Gareta D, et al. Trends in population HIV viral suppression: a longitudinal analysis. AIDS. 2025;39: 1088–1092. doi:10.1097/QAD.0000000000004183

9. Thosago MJ, Lowane MP. A qualitative exploration of psychosocial determinants influencing viral load suppression among adolescents living with HIV: Application of social action theory. Health SA Gesondheid. 2026;31. doi:10.4102/HSAG.v31i0.3093

10. Venter WDF, Moorhouse M, Sokhela S, Fairlie L, Mashabane N, Masenya M, et al. Dolutegravir plus Two Different Prodrugs of Tenofovir to Treat HIV. New England Journal of Medicine. 2019;381: 803–815. doi:10.1056/NEJMoa1902824

11. Wilson HI, Mapesi H. Rollout of dolutegravir-based antiretroviral therapy in sub-Saharan Africa and its public health implications. Pan African Medical Journal. 2020;37. doi:10.11604/pamj.2020.37.243.25512

12. Long L, Kuchukhidze S, Pascoe S, Nichols BE, Fox MP, Cele R, et al. Retention in care and viral suppression in differentiated service delivery models for HIV treatment delivery in sub Saharan Africa: a rapid systematic review. J Int AIDS Soc. 2020;23. doi:10.1002/jia2.25640

13. Kippen A, Nzimande L, Gareta D, Iwuji C. The viral load monitoring cascade in HIV treatment programmes in sub-Saharan Africa: a systematic review. BMC Public Health. 2024;24: 2603. doi:10.1186/s12889-024-20013-x

14. Mugglin C, Kläger D, Gueler A, Vanobberghen F, Rice B, Egger M. The HIV care cascade in sub Saharan Africa: systematic review of published criteria and definitions. J Int AIDS Soc. 2021;24. doi:10.1002/jia2.25761

15. Kamori D, Barabona G, Maokola W, Rugemalila J, Mahiti M, Mizinduko M, et al. HIV viral suppression in the era of dolutegravir use: Findings from a national survey in Tanzania. Onwuamah CK, editor. PLoS One. 2024;19: e0307003. doi:10.1371/journal.pone.0307003

16. Hladik W, Stupp P, McCracken SD, Justman J, Ndongmo C, Shang J, et al. The epidemiology of HIV population viral load in twelve sub-Saharan African countries. Ortega JA, editor. PLoS One. 2023;18: e0275560. doi:10.1371/journal.pone.0275560

17. Gandla S, Nakka R, Khan RA, Bose E, Ghebremichael M. Biological and Social Predictors of HIV-1 RNA Viral Suppression in ART Treated PWLH in Sub-Saharan Africa. Trop Med Infect Dis. 2025;10: 24. doi:10.3390/tropicalmed10010024

18. Fornah L, Shimbre MS, Osborne A, Ma W. Trends and spatial patterns of human immunodeficiency virus prevalence among men and women aged 15–49 in Sierra Leone: evidence from the 2008, 2013 and 2019 Demographic and Health Surveys. BMJ Glob Health. 2025;10: e017208. doi:10.1136/bmjgh-2024-017208

19. Getnet M, Bitew DA, Maru L, Tesfaye E, Adugna DG. Spatial variation of HIV testing and associated factors among pregnant women: a Spatial and multilevel analysis, DHS of sub-Saharan African countries. BMC Public Health. 2025;25: 1375. doi:10.1186/s12889-025-22614-6

20. Mwakyomo J, Sangeda RZ, Mushi H, Njau P. Time to registry discontinuity in Tanzania’s national HIV care registry: a survival analysis of population mobility patterns. 2026. pp. 1–23. doi:10.64898/2026.03.07.26347830

21. Lugoba MD, Sangeda RZ, De Vrieze L, Mushi H, Mutagonda RF, Mwakyomo J, et al. Geographic variation in loss to follow-up from HIV care in Tanzania and its association with pharmacy refill adherence in routine programme data. 2026. doi:10.64898/2026.03.04.26347648

22. Manyanga VP, Mwakyomo J, Mushi H, Lugoba MD, Sambu V, Mutagonda RF, et al. Geospatial Quantification of Antiretroviral Therapy Consumption in Tanzania (2017-2021) Using the WHO Defined Daily Dose Methodology. 2025. doi:10.64898/2025.12.06.25341745

23. Rugemalila J, Kunambi PP, Amour M, Sambu V, Kisonjela F, Rugarabamu A, et al. Trends and correlates in HIV viral load monitoring and viral suppression among adolescents and young adults in Dar es Salaam, Tanzania. Tropical Medicine & International Health. 2024;29: 792–800. doi:10.1111/tmi.14031

24. Rakhmanina N, Foster C, Agwu A. Adolescents and young adults with HIV and unsuppressed viral load: where do we go from here? Curr Opin HIV AIDS. 2024;19: 368–376. doi:10.1097/COH.0000000000000880

25. Asare K, Sookrajh Y, van der Molen J, Khubone T, Lewis L, Lessells RJ, et al. Clinical outcomes with second-line dolutegravir in people with virological failure on first-line non-nucleoside reverse transcriptase inhibitor-based regimens in South Africa: a retrospective cohort study. Lancet Glob Health. 2024;12: e282–e291. doi:10.1016/S2214-109X(23)00516-8

26. Meque I, Herrera N, Nhangave A, Mandlate D, Guilaze R, Tambo A, et al. The rollout of paediatric dolutegravir and virological outcomes among children living with HIV in Mozambique. South Afr J HIV Med. 2024;25. doi:10.4102/sajhivmed.v25i1.1578

27. Mushi H, Lugoba MD, Sangeda RZ, Mutagonda RF, Mwakyomo J, Musiba G, et al. Predictors of loss to follow-up among patients receiving antiretroviral therapy in Njombe Region, Tanzania, 2017–2021. 2026. doi:10.64898/2026.02.28.26347333

28. Kalulo MB, Sangeda RZ, Mwakyomo J, Sangeda GR, Sambu V, Njau P. Utilizing pharmacy refill data to predict loss to follow-up among people living with HIV in Manyara region of Tanzania. 2026. doi:10.64898/2026.02.24.26347034

29. Lugoba MD, Sangeda RZ, Mutagonda RF, Mwakyomo J, Musiba G, Sambu V, et al. Leveraging Machine Learning Models and Pharmacy Refill Adherence as a Cost-Effective Proxy for Predicting HIV Viral Suppression during Antiretroviral Therapy in Resource-Limited Settings. 2026. doi:10.64898/2026.01.05.26343496

